# Challenges in the case-based surveillance of infectious diseases

**DOI:** 10.1101/2023.12.19.23300224

**Authors:** Oliver Eales, James M. McCaw, Freya M. Shearer

## Abstract

To effectively inform infectious disease control strategies, accurate knowledge of the pathogen’s transmission dynamics is required. The infection incidence, which describes the number of new infections in a given time interval (e.g., per day or per week), is fundamental to understanding transmission dynamics, and can be used to estimate the time-varying reproduction number and the severity (e.g., the infection fatality ratio) of a disease. The timings of infections are rarely known and so estimates of the infection incidence often rely on measurements of other quantities amenable to surveillance. Case-based surveillance, in which infected individuals are identified by a positive test, is the predominant form of surveillance for many pathogens, and was used extensively during the COVID-19 pandemic. However, there can be many biases present in case-based surveillance indicators due to, for example, test sensitivity and specificity, changing testing behaviours, and the co-circulation of pathogens with similar symptom profiles. Without a full understanding of the process by which surveillance systems generate data, robust estimates of the infection incidence, time-varying reproduction number, and severity based on these data cannot be made. Here we develop a mathematical description of case-based surveillance of infectious diseases. By considering realistic epidemiological parameters and situations, we demonstrate potential biases in common surveillance indicators based on case-based surveillance data. The description is highly general and could be applied to a diverse set of pathogens and situations. The mathematical description could be used to inform inference of infection incidence using existing data, with a full understanding of where bias and uncertainty will be present in any such analysis. Future surveillance strategies could be designed to minimise these sources of bias and uncertainty, providing more accurate estimates of a pathogen’s transmission dynamics and, ultimately, more targeted application of public health measures.

## 1 Introduction

Diseases caused by the spread of infectious pathogens result in substantial mortality and morbidity. There is a highly diverse set of pathogens circulating worldwide [1], with varying levels of clinical severity, geographic distribution, and infection prevalence. To effectively inform public health strategies and interventions, accurate knowledge of the infection rates of a pathogen, and how they change over time are required.

The infection incidence of a pathogen describes the number of individuals infected with the pathogen in a given time interval (e.g., per day or per week). Estimates of the infection incidence provide information on the total burden of infection, which is important for estimating the severity of the pathogen (i.e., the infection fatality ratio and infection hospitalisation ratio). The time series of infection incidence is also intrinsically linked to the time-varying reproduction number, *R*(*t*) [2], which details the expected number of secondary infections an infected individual will generate [3]; *R*(*t*) is routinely estimated during epidemics and pandemics to better inform predictions of future levels of infection and so improve public health strategies [4, 5].

Measuring the timing of all infections is in practice impossible and so the infection incidence is never known. Instead, data are collected for other downstream points of the infection process (e.g., symptom onset, hospitalisation, death). A frequently made (implicit) assumption is that these epidemic time series data are a reliable proxy for the infection incidence [6]; estimates of *R*(*t*) made under this assumption (using these epidemic time series data) can be heavily biased, particularly during periods in which *R*(*t*) varies rapidly [7]. The relationship between the infection incidence and these epidemic time series needs to be well described for the outputs of such inference to be safely interpreted.

Case-based surveillance relies on the identification of persons infected with the pathogen (cases) [8]. During the COVID-19 pandemic, cases of COVID-19 were primarily identified through mass testing programs where individuals could seek a test through community-based testing centres. In some countries, cases were also identified through more targeted testing of the population. For example, in England the REACT-1 [9] and ONS CIS [10] studies tested random subsets of the population (chosen to be representative of the entire population), to estimate the infection prevalence of SARS-CoV-2 over time. For other pathogens (and for the continuing surveillance of COVID-19), cases are primarily identified through testing performed at health-care services, and so case time-series are dependent on healthcare seeking behaviour [11, 12].

The relationship between the daily number of cases (i.e, identified infections) and infection incidence can be complex, depending on a myriad of factors — for example, the symptomatic proportion, test sensitivity, and health-care and test-seeking behaviours — some of which may change over time. When using data generated by case-based surveillance to determine transmission dynamics, these relationships must be well understood and accounted for. Improved characterisation of the processes by which individuals are identified by case-based surveillance systems is required to interpret existing data, and to optimise the design of future case-based surveillance strategies. While studies have previously described aspects of these processes [13, 14, 15, 16], the exact form of the data-generating process has not been investigated.

Here we develop a mathematical description of case-based surveillance and use it to demonstrate challenges associated with the surveillance of pathogens. The mathematical description is highly general, and could be used to inform surveillance strategies for many different pathogens in diverse settings. Though our description is mathematical by necessity, we discuss implications and future considerations for surveillance with a general audience in mind.

## 2 A mathematical description of case-based surveillance

### 2.1 The proportion of the population exhibiting symptoms

For a given disease, the probability of an individual taking a test at any time is dependent on their symptom status at that time. For simplicity, we assume a binary symptom status; individuals either exhibit (disease-specific) symptoms or they do not. This binary variable could be defined for any specific, or non-specific symptom set. Following infection with the disease, the probability of the infected individual exhibiting symptoms is *F* (*τ*), a function of the time since infection, *τ*.

In general *F* (*τ*) will be composed of two other functions (Fig 1). The first function, *f*_1_(*τ*), describes the probability of symptom onset as a function of time since infection, *τ*. The sum total of this probability distribution, ∫ *f*_1_(*τ*), will equal the total proportion of infections that go on to exhibit symptoms (which can be less than one when asymptomatic infections occur). The second function, *f*_2_(*θ*) describes the probability of exhibiting symptoms as a function of time since symptom onset, *θ*. Note that this function can also be written in terms of the probability distribution function for symptom duration (one minus the cumulative distribution function). *F* (*τ*) can then be written in terms of these two composite functions as:

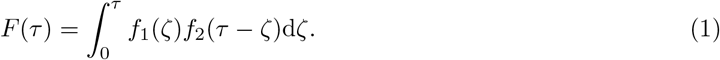

If the infection incidence (the proportion of the population infected at time *t*) is described by *I*(*t*) then the proportion of the population exhibiting symptoms due to infection (with the pathogen of interest) at time *t* is given by:

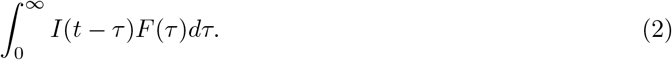

Note that this assumes that the time between successive infections for any individual is larger than the time taken for *F* (*τ*) to go to zero. In other words, this expression for the proportion of the population exhibiting symptoms due to infection will not count any individuals more than once. If we assume that there is another underlying process in the population (either a contagious disease or otherwise, but unrelated to the pathogen of interest) resulting in a proportion, *S*(*t*), of the population exhibiting symptoms at time *t*, then the proportion of the population exhibiting symptoms, *n*_*s*_(*t*), is now given by:

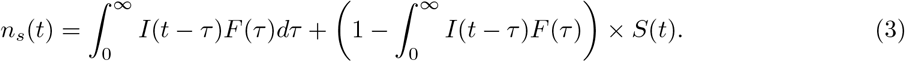

Similarly the proportion not exhibiting symptoms is given by:

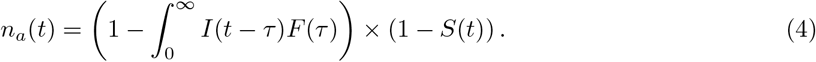

A visualisation of the paper’s entire (see below) mathematical description is provided in figure 2.

**Figure 1.**
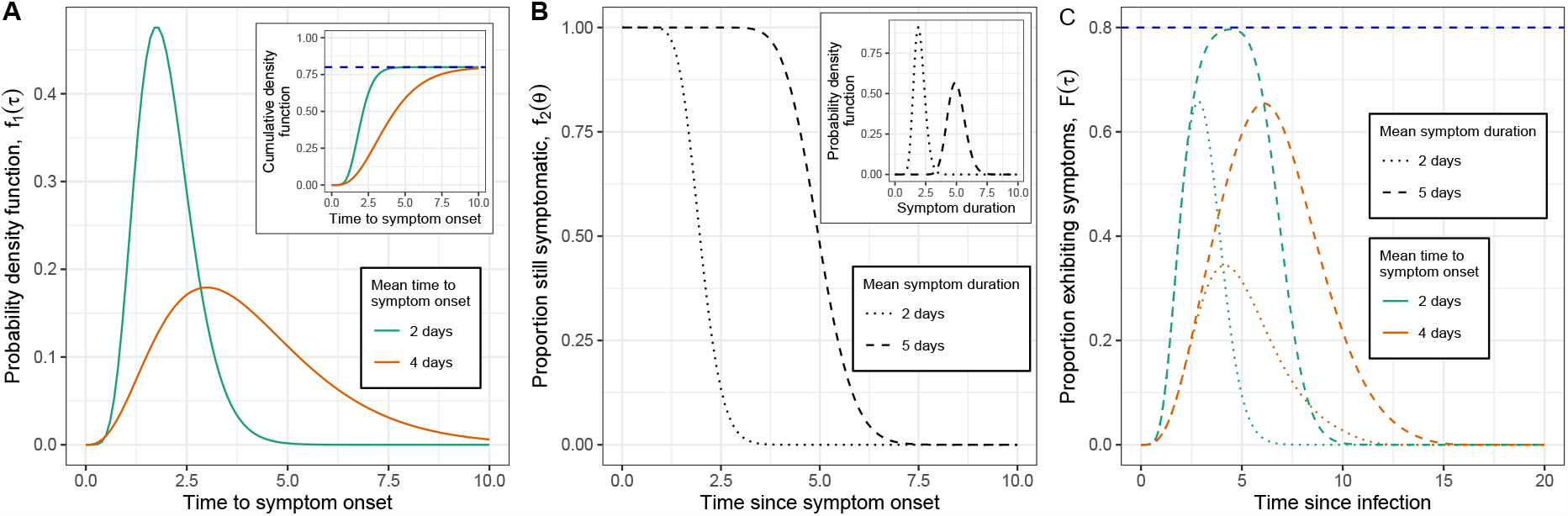
Illustrative example distributions of *F* (*τ*) for different valued distributions of *f*_1_(*τ*) and *f*_2_(*θ*). (A) The probability density function of the time to symptom onset (following infection), *f*_1_(*τ*) for two example distributions. Both example distributions were gamma distributed, with means of 4 days (orange, shape parameter = 4, rate parameter = 1) and 2 days (green, shape parameter = 8, rate parameter = 4). The total proportion of infections exhibiting symptoms is 0.80 for both distributions and is demonstrated in the inset plot of the cumulative distribution function (dashed blue line). (B) The proportion of the population exhibiting symptoms as a function of time since symptom onset, *f*_2_(*θ*), for two example distributions. Both example distributions were calculated as one minus the cumulative density function of gamma distributions describing symptom duration (inset). The gamma distributed functions have means of 2 days (dotted line, shape parameter = 20, rate parameter = 10), and 5 days (dashed line, shape parameter = 50, rate parameter = 10). (C) The proportion of infections exhibiting symptoms as a function of time since infection, *F* (*τ*), for all four combinations of the example distribution of *f*_1_(*τ*) and *f*_2_(*τ*) given in (A) and (B).

**Figure 2.**
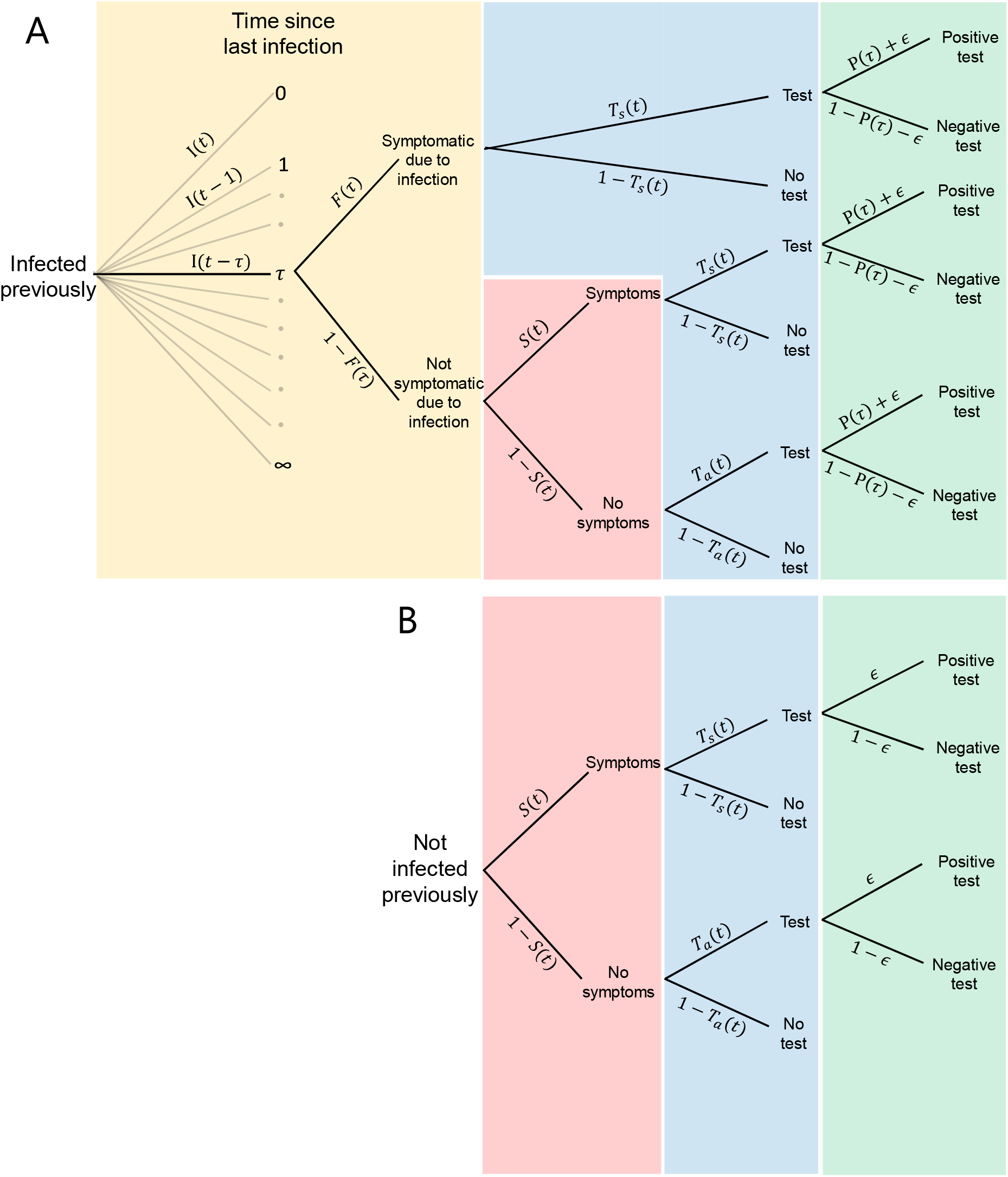
A graphical representation of the mathematical description of case-based surveillance. (A,B) The probability tree showing the probability of possible outcomes of all individuals in a population. (A) The probability tree for individuals who have been infected at some point in the past. The tree is shown for a single value of time since infection, *τ*, but in general, there is an equivalent tree for all values of *τ*. The probability distribution of the time since last infection (for individuals infected previously) is given by the incidence of infection at previous times, *I*(*t −τ*); these infected individuals will exhibit symptoms due to their infection with a probability *F* (*τ*) (orange shaded region). Those who are not symptomatic due to infection will have a probability of exhibiting symptoms (due to background symptom rates in the population) as a function of time, *S*(*t*) (red shaded region). The probability of taking a test is time-dependent and takes a value of *T*_*s*_(*t*) for symptomatic individuals and *T*_*a*_(*t*) for asymptomatic individuals (blue shaded region). Finally for those who take a test their probability of testing positive is a function of the time since infection, *P* (*τ*) + *ϵ*. (B) The probability tree for individuals who have not been infected at any point. This tree is analogous to the tree for those infected a long time ago (as *τ → ∞* : *F* (*τ*) *→* 0, *P* (*τ*) *→* 0).

### 2.2 The rate of symptomatic and asymptomatic testing

We assume that individuals who exhibit symptoms have a daily testing probability of *T*_*s*_(*t*), and those who do not exhibit symptoms have a daily testing probability of *T*_*a*_(*t*). The daily total rate of testing, *N* (*t*), can be written in terms of its symptomatic, *N*_*s*_(*t*), and asymptomatic, *N*_*a*_(*t*) components:

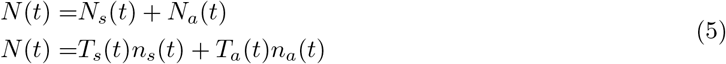

where the first term represents symptomatic testing and the second term represents asymptomatic testing. Note that more generally the daily testing probability for an individual exhibiting symptoms could also be dependent on the time since the onset of symptoms (e.g., an individual may be more likely to test when they first exhibit symptoms). The equations could be extended in a straightforward manner to include this additional complexity, but this is not required for the effects we explore in this paper.

### 2.3 The probability of testing positive when tested

Having only described the number of tests that will be performed, we now describe the number of positive tests that will be obtained. In general, the probability an individual tests positive will be a function of the time since their last infection, *τ*. Individuals who have never been infected (or who were infected a long-time ago) will also have a small probability of testing positive due to imperfect specificity of the test. We model the probability of testing positive (Fig 3) as

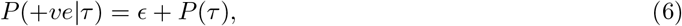

where *ϵ* is a small constant probability of testing positive due to imperfect specificity, and *P* (*τ*) describes the increased probability of testing positive following infection and tends to zero for large *τ*. Note that *P* (*τ*) + *ϵ <* 1 for all values of *τ*.

**Figure 3.**
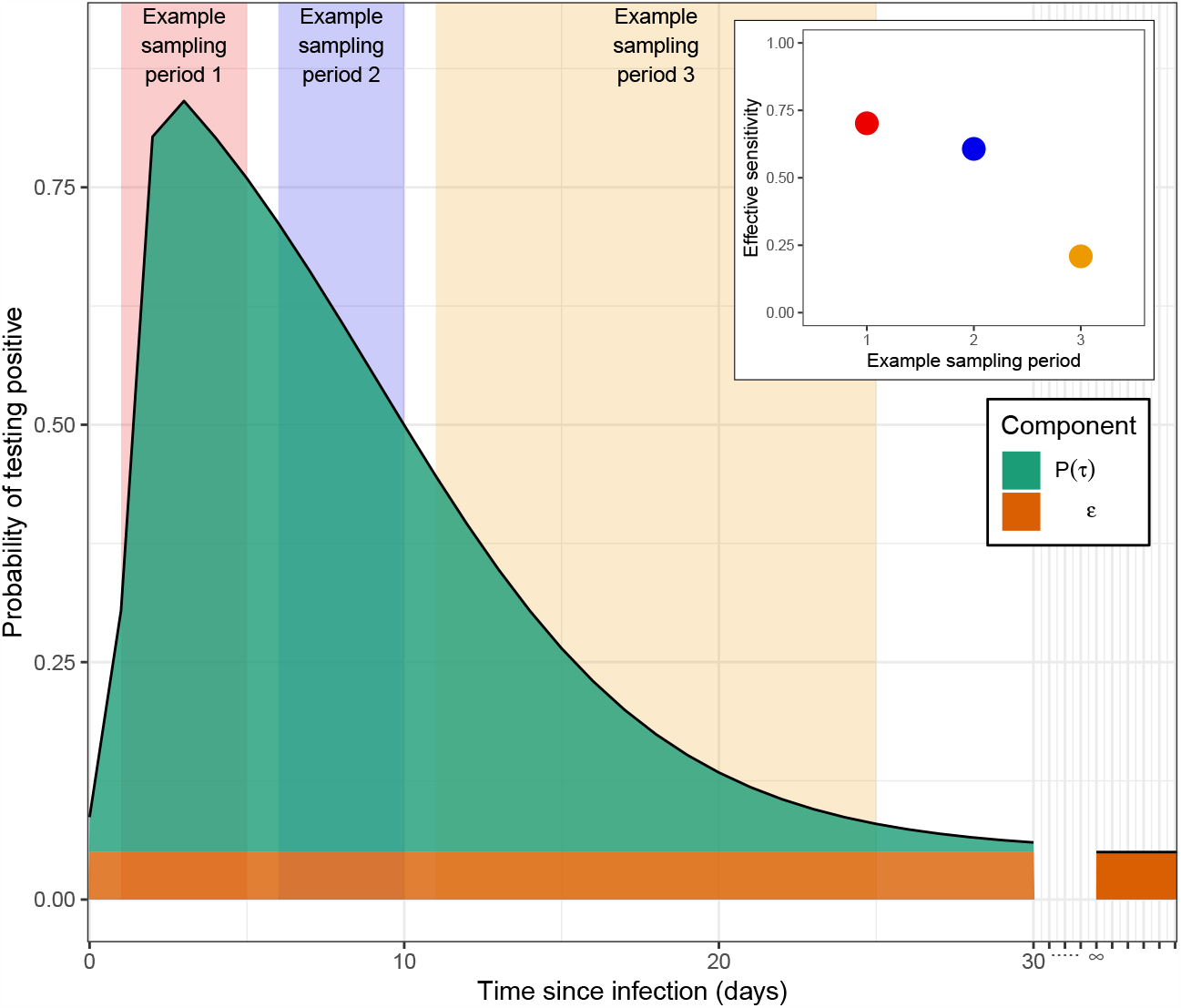
An example distribution for the probability of testing positive as a function of time since infection for an individual. The probability is composed of two components: *ϵ*, the probability of testing positive irrespective of infection (e.g., when not previously infected or when infected a long time ago) (orange); and *P* (*τ*), the increased probability of testing positive following infection, which tends to zero for long times since infection. *P* (*τ*) in this figure is plotted at discrete time points (days), and takes the form *logit*^*−*1^ (*β*_1_ + *β*_2_(*τ − C*) + *δ*_*τ−C>*0_ *× β*_2_*β*_3_(*τ − C*)) where *δ*_*τ−C>*0_ is 1 if *τ − C >* 0 and 0 if not [17]. The parameters used were: *β*_0_ = 1.51, *β*_1_ = 2.19, *β*_2_ = *−*1.1 and *C* = 2.18. Also shown is the effective sensitivity (inset plot) of the test if all sampling was performed at a constant rate in the three example sampling periods shown (shaded rectangles).

### 2.4 The symptomatic and asymptomatic case rate

The daily proportion of the population identified as a positive case, *C*(*t*) (including those with and without the infection), can be written in terms of a symptomatic component, *C*_*s*_(*t*), and asymptomatic component, *C*_*a*_(*t*):

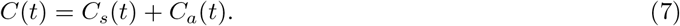

The symptomatic component can be written as

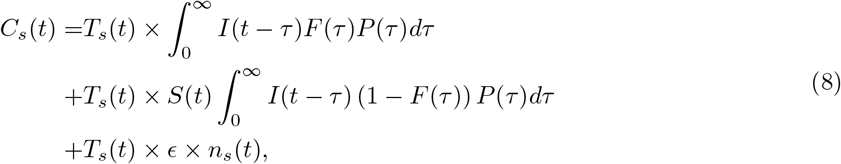

and the asymptomatic component can be written as

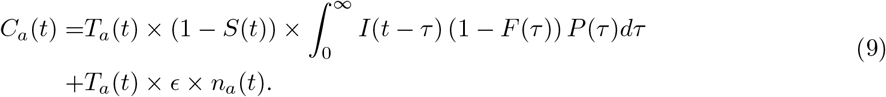

The symptomatic component consists of three terms (equation 8), all of which arise due to testing individuals who are exhibiting symptoms. The first term represents cases where symptoms have been directly caused by infection. The second term represents cases where, at the time of testing, there were no symptoms due to infection, but there were unrelated symptoms. The final term represents the cases that would have been identified (at the overall symptomatic testing rate, *n*_*s*_(*t*)*T*_*s*_(*t*)) irrespective of infection incidence, due to imperfect test specificity. The asymptomatic component consists of two terms (equation 9), all of which arise due to testing individuals who are not exhibiting symptoms. The first term represents cases in which there are no symptoms caused by infection (no symptoms at the time of testing) and for which no background symptoms occur. The second term represents the cases that would have been identified (at the overall asymptomatic testing rate, *n*_*a*_(*t*)*T*_*a*_(*t*)) irrespective of infection incidence, due to imperfect test specificity. A visualisation of this mathematical description is provided in figure 2. Note that throughout the mathematical description presented in this paper, asymptomatic cases, *C*_*a*_(*t*), includes asymptomatic, pre-symptomatic and post-symptomatic infections.

## 3 Implications for surveillance

In this section, we consider a range of pathogen characteristics and surveillance approaches and use the mathematical expressions above to highlight key challenges and opportunities in the surveillance of infectious diseases. The daily proportion of the population identified as a positive case, *C*(*t*), is composed of a symptomatic and asymptomatic component. The symptomatic and asymptomatic testing probabilities, *T*_*s*_(*t*) and *T*_*a*_(*t*), effectively scale these components. In general, *T*_*s*_(*t*) and *T*_*a*_(*t*) will have different values — there will most likely be a higher probability of testing when symptomatic — and *C*(*t*) will be dependent on numerous distributions and testing probabilities (equations 7, 8 and 9).

### 3.1 Random testing

In the special case when *T*_*s*_(*t*) and *T*_*a*_(*t*) are equal to some testing rate *T* (*t*), *T* (*t*) = *T*_*s*_(*t*) = *T*_*a*_(*t*), then equations 7, 8 and 9 simplify to:

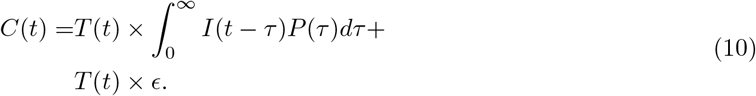

This special instance describes the infection prevalence of the pathogen,

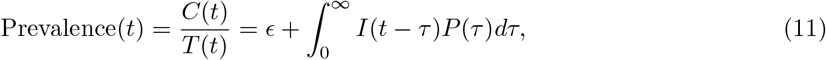

and shows that the infection prevalence does not depend on the rate of symptoms (symptoms caused by infection, *F* (*τ*), or the background rate of symptoms, *S*(*t*)). Infection prevalence can be measured using random sampling, which ensures that the probability of testing is the same irrespective of symptom status. As can be seen from equation 11, if the time series of infection prevalence is measured, then only knowledge of how test positivity varies over time since infection (*P* (*τ*) and *ϵ*) is required to estimate the time series of infection incidence.

### 3.2 Test timing and accuracy

The accuracy of a test that reports on the presence/absence of a pathogen is often described in terms the test’s sensitivity and specificity. Sensitivity is the probability of testing positive for an infected individual. Specificity is the probability of testing negative for an uninfected individual. However, the probability of a positive test following infection typically depends on the time since infection. Estimates of the sensitivity and specificity of a test are thus dependent on when infected individuals are tested. Additionally, estimates of the sensitivity and specificity will depend on for how long following infection an individual is classified as infected — this classification may not be consistent across locations and time periods.

Estimates of a test’s sensitivity depends on how soon following infection infected individuals are tested on average. This ‘effective sensitivity’ will be different for different testing strategies (Fig 3). For example: testing contacts of infected individuals (identified through contact tracing) could result in a small average time between infection and test; symptomatic testing could result in an average time between infection and test approximately matching the time between infection and symptom onset (with a possible delay from symptom onset to test); and testing upon hospitalisation could result in an even longer delay between infection and test. If testing is performed at a rate, *ψ*(*τ*), then the effective sensitivity will be:

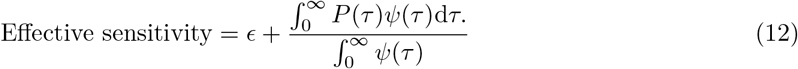

Note that this assumes that an infected individual is classified as infected until (at least) *P* (*τ*) goes to 0. Under this assumption, the test’s specificity is simply given by:

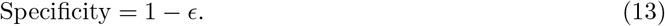

If infected individuals are classified as individuals infected within a specific time period (e.g., less than *τ*_*i*_ days since infection), and *P* (*τ*) does not go to zero by the end of this period (*P* (*τ*_*i*_) *>* 0) then the expression for estimates of sensitivity and specificity would need to be adjusted as follows. For estimates of the test’s sensitivity, the sampling rate, *ψ*(*τ*), would now only consider individuals infected within the given period (*τ < τ*_*i*_). Estimates of the test’s specificity would now be time-dependent due to a dependence on the incidence of infection; infected individuals will have an increased probability of testing positive even after they are no longer classified as infected (the population of ‘uninfected’ individuals will now include some of these recently infected individuals).

### 3.3 The asymptomatic to symptomatic case ratio

In general the number of asymptomatic and symptomatic cases will depend on the background proportion symptomatic, *S*(*t*), and how imperfect test specificity is, *ϵ* (see Sections 3.4–3.6). However, we first consider the simple scenario in which *S*(*t*) = 0 and *ϵ* = 0. Under these assumptions the ratio of symptomatic cases to asymptomatic cases is given by (see Box 1):

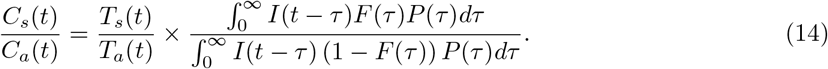

The ratio is firstly a function of the relative testing probabilities (left-hand side of expression). These testing probabilities can vary over time. For example, test-seeking behaviour, clinical decision making, testing strategies, and testing capacities can change, influencing these probabilities. When the symptomatic testing probability increases (relative to the asymptomatic testing probability) the symptomatic case proportion will increase (and vice versa). Even when the relative testing probabilities are constant (or known and adjusted for) the ratio of symptomatic to asymptomatic cases will, in general, vary over time (Figure 4A) due to changes in the infection incidence (right-hand side of expression). The ratio will only be constant if the infection incidence, *I*(*t*), is constant, or the distributions describing positivity, *P* (*τ*), and symptom status, *F* (*τ*), are in a form such that the ratio of integral terms in the expression are constant.

**Figure 4.**
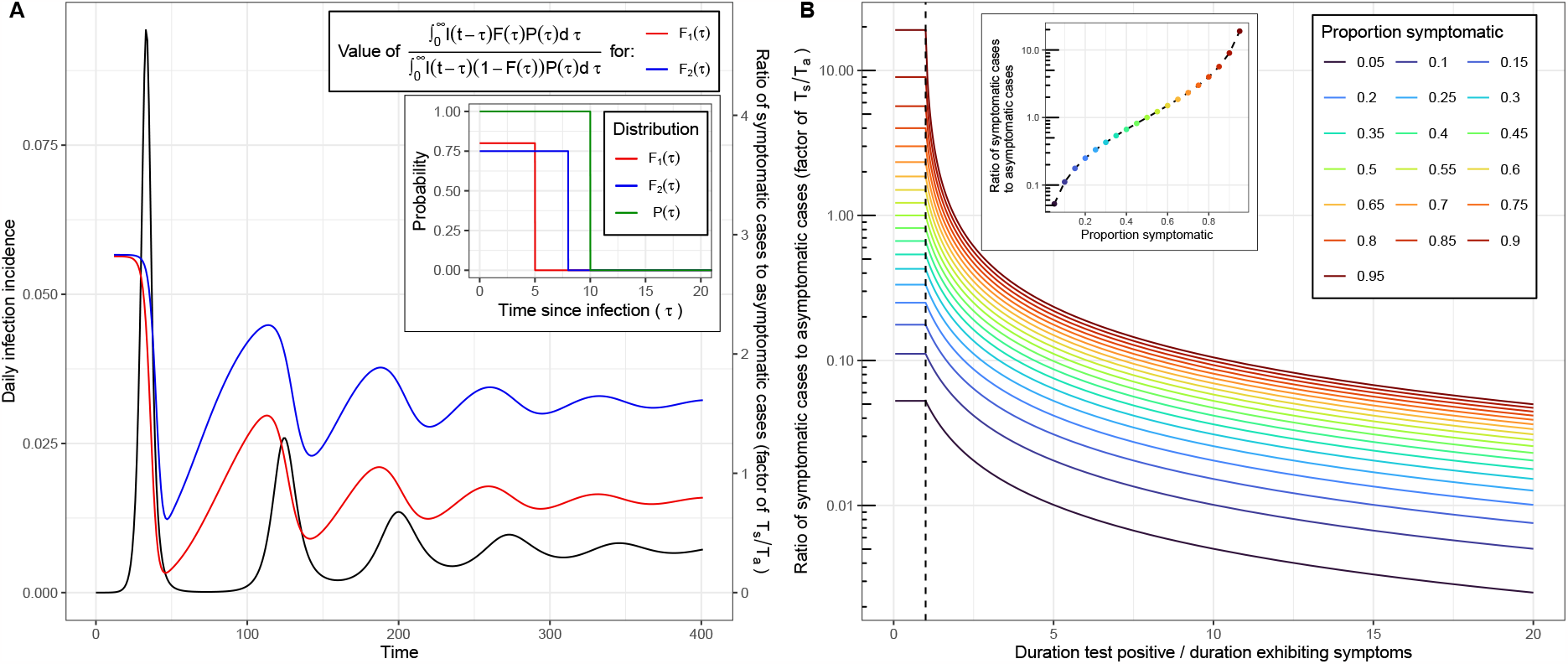
The relative magnitude of symptomatic and asymptomatic cases. (A) The ratio of symptomatic cases and asymptomatic cases (assuming *S*(*t*) = 0 and *ϵ* = 0), as a factor of their relative testing rates (*T*_*s*_(*t*)*/T*_*a*_(*t*)) (colored lines, right-hand y-axis, equation given), for an example function of daily infection incidence (black line, left-hand y-axis) simulated using an SIRS model [18]. The distributions for *F* (*τ*) (inset) were constant with value *h*_*f*_ for *τ* ≤ *τ*_*f*_ and 0 otherwise (see Box 1). Similarly, the distributions for *P* (*τ*) (inset) was constant with value *h*_*p*_ for *τ* ≤ *τ*_*p*_ and 0 otherwise (see Box 1). A single distribution was used for *P* (*τ*) (green, *h*_*p*_ = 1, *τ*_*p*_ = 10) and two distributions were used for *F* (*τ*), *F*_1_(*τ*) (red, *h*_*f*_ = 0.8, *τ*_*f*_ = 5) and *F*_2_(*τ*) (blue, *h*_*f*_ = 0.75, *τ*_*f*_ = 8). (B) The ratio of symptomatic and asymptomatic cases, as a factor of their relative testing rates (*T*_*s*_(*t*)*/T*_*a*_(*t*)), as a function of the duration an infected individual tests positive (*τ*_*p*_) divided by the duration a symptomatic individual exhibits symptoms (*τ*_*f*_) (solid lines). Different coloured lines are shown for different values of the peak proportion symptomatic (*h*_*f*_). The ratio of symptomatic to asymptomatic cases is shown as a function of the peak proportion symptomatic (inset) for a duration of testing positive equal to the duration of symptoms (*τ*_*f*_ = *τ*_*p*_) (black dashed vertical line passing through 1 on the x-axis).

The relative magnitude of symptomatic and asymptomatic cases is heavily dependent on the distributions describing positivity and symptom status (Box 1), and their overlap. In general, the greater the proportion of infections that go on to exhibit symptoms, and the greater the duration of symptoms, the greater the ratio of symptomatic to asymptomatic cases (Figure 4B). This ratio is at its greatest when individuals only test positive during the period in which they are likely to exhibit symptoms. When the typical duration for which an infected individual tests positive increases past the typical period in which they exhibit symptoms (if they exhibit symptoms), the ratio of symptomatic cases to asymptomatic cases begins to decrease. If infected individuals test positive for far longer than the duration of symptoms, and the asymptomatic and symptomatic testing probabilities are similar, the symptomatic proportion of cases would be small.

#### Box 1

**Ratio of asymptomatic and symptomatic cases**

Under the condition that *S*(*t*) = 0 and *ϵ* = 0 (assumed throughout Box 1), the ratio of symptomatic and asymptomatic cases is given by the ratio of the first expressions in equations 8 and 9:

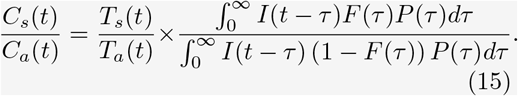

In general this expression will vary over time due to the dependence on the ratio of symptomatic and asymptomatic testing probabilities (*T*_*s*_(*t*)*/T*_*a*_(*t*)), and on the incidence (*I*(*t*)). When *I*(*t*) is constant the expression can be rewritten as:

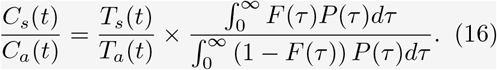

The relative magnitude of symptomatic and asymptomatic cases is now represented by the ratio of testing probabilities multiplied by a constant. The constant is determined by the distributions, *P* (*τ*) and *F* (*τ*). Let us assume both distributions follow a top-hat function with:

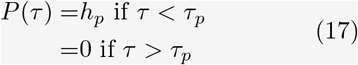

and

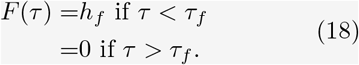

Here *h*_*f*_ is the proportion of infected individuals who exhibit symptoms and all symptoms begin at the time of infection. The parameter *τ*_*f*_ is the duration of symptoms and is the same for all individuals that exhibit symptoms. Similarly, *h*_*p*_ is the probability of a recently infected individual testing positive and *τ*_*p*_ is the duration for which they will test positive (with probability *h*_*p*_). With these distributions for *F* (*τ*) and *P* (*τ*), the expression for the ratio of symptomatic and asymptomatic cases depends on the relative value of *τ*_*f*_ and *τ*_*k*_. When *τ*_*f*_ *> τ*_*k*_:

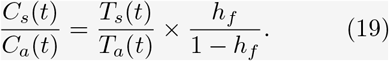

When *τ*_*f*_ *< τ*_*k*_:

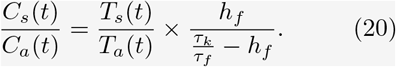

The longer the duration an individual tests positive following infection relative to the duration of symptoms, the greater the magnitude of asymptomatic cases relative to symptomatic cases. If individuals exhibit symptoms over the entire period they test positive (*τ*_*f*_ *> τ*_*k*_) then the ratio of symptomatic to asymptomatic cases reaches its highest value and is determined by the proportion of infections that are symptomatic.

### 3.4 Co-circulating pathogens with similar symptom profiles

We explored the impact of the background proportion of individuals exhibiting symptoms, *S*(*t*) (assumed constant in time to begin with), on four different surveillance indicators: the proportion of the population exhibiting symptoms (i.e., as measured through syndromic surveillance or symptom surveys); symptomatic cases (identified through symptomatic testing); asymptomatic cases (identified through asymptomatic testing); and all cases identified through mass testing (i.e., the linear combination of symptomatic and asymptomatic cases) (Figure 5). As one would expect, a larger value for *S*(*t*) means a greater proportion of the population exhibit symptoms. Increasing *S*(*t*) increased the number of symptomatic cases, and decreased the number of asymptomatic cases, since these asymptomatic cases are exhibiting symptoms unrelated to their infection status. For mass testing, a larger (but constant) *S*(*t*) led to either an increase (*T*_*s*_(*t*) *> T*_*a*_(*t*)) or decrease (*T*_*s*_(*t*) *< T*_*a*_(*t*)) in the number of cases depending on the relative testing probabilities; when testing probabilities were equal there was no change, as expected (see Section 3.1). Even under the assumption of a constant value of *S*(*t*) the changes (multiplicative and additive) in the four surveillance indicators were not constant over time (Supplementary Figures S1 and S2), due to their dependence on infection incidence, which in general is not constant. The only exception was for asymptomatic testing in which the multiplicative change was constant. The additive change for the proportion symptomatic was only approximately constant for small values of *S*(*t*).

**Figure 5.**
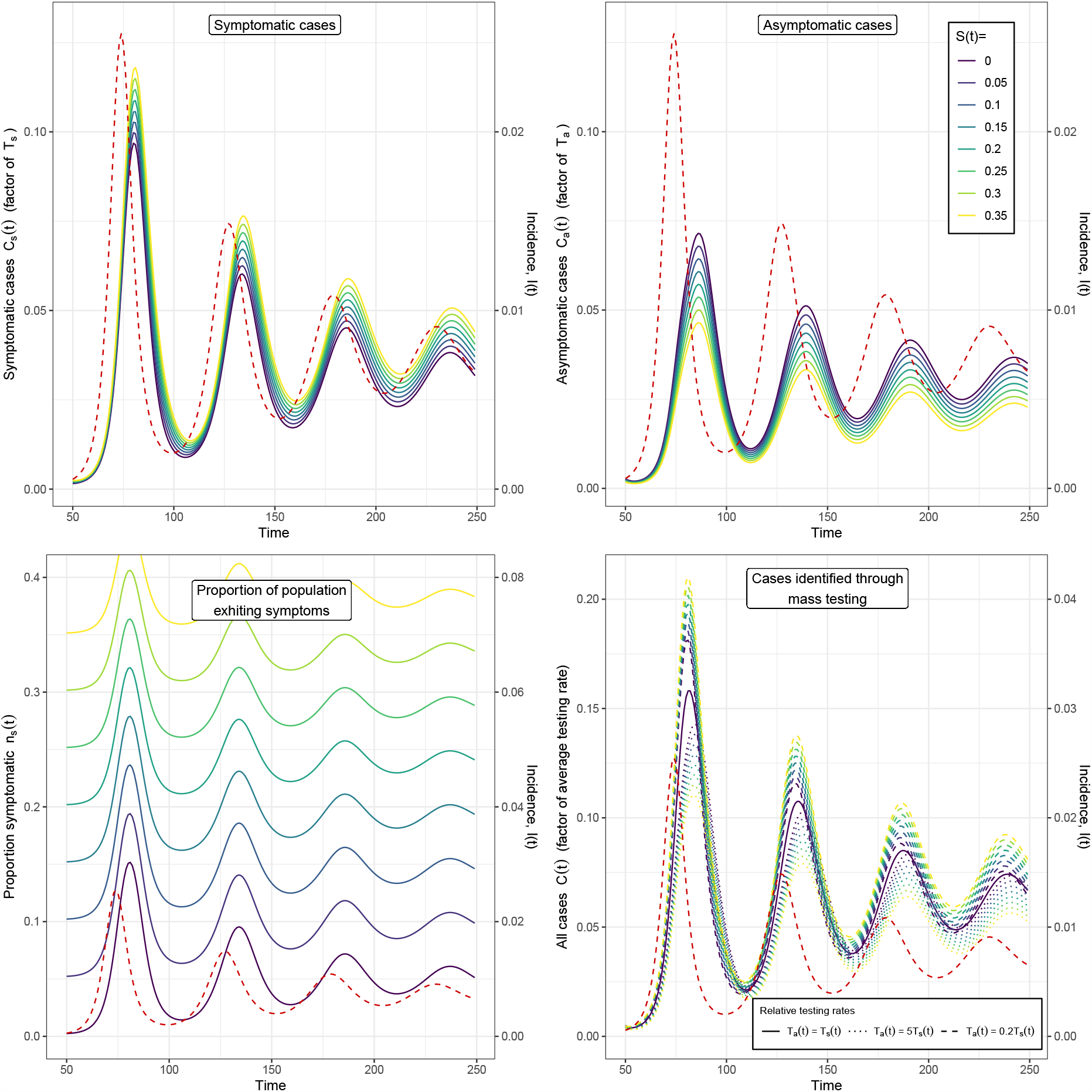
The effect of changing *S*(*t*) on the time series of surveillance indicators. The time series of four possible surveillance indicators (left y-axis): symptomatic cases; asymptomatic cases; proportion of the population exhibiting symptoms; and all cases identified through mass testing (combined total of symptomatic and asymptomatic cases). The time series for the surveillance indicators were calculated assuming different constant values of *S*(*t*) (coloured lines, legend in top-right) for a single example time series of infection incidence (red dashed line, right y-axis) which was simulated using an SIRS model [18]. The distribution of *F* (*τ*) was composed of *f*_1_(*τ*) and *f*_2_(*θ*) (see Section 2.1). *f*_1_(*τ*) was assumed to be gamma distributed with a mean of 3 days (shape parameter = 9, rate parameter = 3). *f*_2_(*θ*) was assumed to be one minus the cumulative distribution function of a gamma distribution with a mean of 8 days (shape parameter = 40, rate parameter = 5). The distribution of *P* (*τ*) was chosen to match an empirical distribution for SARS-CoV-2 PCR positivity [17], with *ϵ* assumed to be zero. Cases identified through mass testing were shown for different relative testing rates: *T*_*a*_(*t*) = *T*_*s*_(*t*) (random testing); *T*_*a*_(*t*) = 5*T*_*s*_(*t*) (higher testing probability for asymptomatic individuals); and *T*_*a*_(*t*) = 0.2*T*_*s*_(*t*) (higher testing probability for symptomatic individuals). Note that for random testing the time series of all cases was the same for all values of *S*(*t*).

The magnitude of effect of changing *S*(*t*) was different for all four surveillance indicators. Changing *S*(*t*) leads to a linear change in all four indicators (Figure 6), with different expressions describing the gradient (Box 2). The expression for the gradient is simple for asymptomatic cases, gradient=-1, which explains the constant multiplicative change over time (when *S*(*t*) is constant). In contrast, the (positive) gradient for symptomatic cases was more complex and was determined by the relative magnitude of symptomatic and asymptomatic cases (assuming equal testing rates) (see Section 3.3). When there are far more symptomatic cases relative to asymptomatic cases, the (positive) gradient will be small. Conversely, when there are far more asymptomatic cases relative to symptomatic cases, the gradient will be larger. For all cases identified through mass testing, the gradient lies somewhere between the individual values for asymptomatic and symptomatic cases, and is zero when the relative testing probabilities are equal.

**Figure 6.**
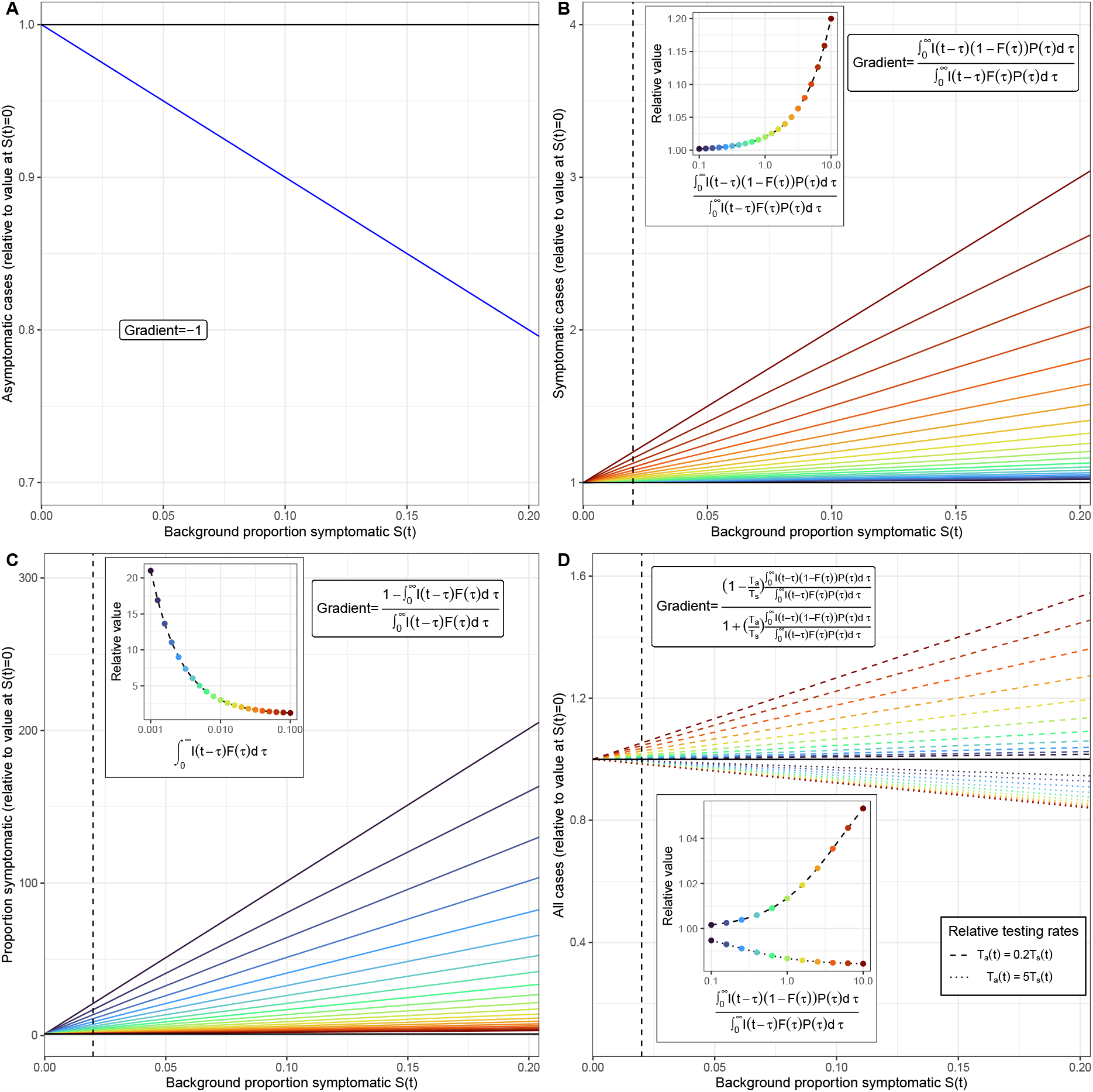
The magnitude of the effect of *S*(*t*) on surveillance indicators. The linear effect of changing the background proportion who are symptomatic, *S*(*t*), (relative to the value at *S*(*t*) = 0) on four different surveillance indicators: (A) asymptomatic cases; (B) symptomatic cases; (C) the proportion of the population exhibiting symptoms; (D) all cases identified through mass testing. For all four surveillance indicators the equation for the gradient of the line is provided (see Box 2). With the exception of the gradient for asymptomatic cases, the gradients are dependent on the infection incidence (and other parameters). The linear effect of changing *S*(*t*) is shown for different values of the integral terms in the gradient equations (coloured lines). The gradient in (B) and (D) is a function of the relative magnitude of symptomatic and asymptomatic cases (assuming equal testing rates) (see Section 3.3). The gradient in (C) is a function of the proportion of the population that are infected and symptomatic due to the infection, 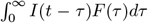. The value of these integral terms are provided in the inset plots (coloured points), as well as their relationship with the linear effect of changing *S*(*t*) (relative to the value at *S*(*t*) = 0) when *S*(*t*) = 0.02 (dashed black line in main and inset plots). All cases identified through mass testing were shown for different relative testing rates: *T*_*a*_(*t*) = 5*T*_*s*_(*t*) (higher testing probability for asymptomatic individuals) (dotted lines); and *T*_*a*_(*t*) = 0.2*T*_*s*_(*t*) (higher testing probability for symptomatic individuals) (dashed lines).

The proportion of the population exhibiting symptoms is the indicator most strongly affected by *S*(*t*). In general the gradient is a function of the proportion of the population that are infected and symptomatic due to the infection. When this value is low, the gradient will be large; for example if only 0.1% of the population are infected and symptomatic due to infection, the gradient will be 999 and the indicator will be significantly altered for even small values of *S*(*t*). Even when the value is relatively high-—could expect a value as high as 10% in certain circumstances (infection levels were very high during the COVID-19 pandemic [19]) — the gradient is still comparatively high (for 10%, gradient would be 9) compared to what might be expected for the other surveillance indicators.

#### Box 2

**Effect of S(t) on surveillance indicators**

A surveillance indicator, *X*(*t*), can be written in terms of its value when *S*(*t*) = 0:

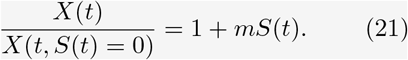

Here *m* is the gradient and describes the effect of *S*(*t*) on *X*(*t*) relative to its value when *S*(*t*) = 0.

**The proportion exhibiting symptoms**

The effect of *S*(*t*) on the proportion of the population exhibiting symptoms can be calculated from equation 3 giving:

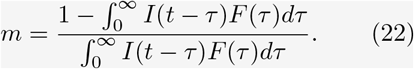

**Asymptomatic cases**

For asymptomatic cases the gradient is trivially obtained from equation 9: *m* = *−*1.

**Symptomatic cases**

For symptomatic cases the gradient is obtained from equation 8 and is given by:

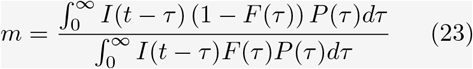

**Total cases**

For the total number of cases, *C*(*t*) = *C*_*a*_(*t*) + *C*_*a*_(*t*), the effect of *S*(*t*) can be calculated from equations 7, 8 and 9:

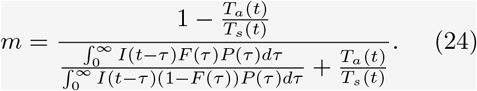

This takes a negative value when *T*_*a*_(*t*) *> T*_*s*_(*t*) and a positive value when *T*_*a*_(*t*) *< T*_*s*_(*t*).

In general, the value of *S*(*t*) will vary over time. For example, another circulating pathogen or allergies (which show seasonal trends) could exhibit similar symptom profiles. The four surveillance indicators would thus be affected by varying amounts over time, depending on the value of *S*(*t*), further complicating interpretation of the quantities illustrated in figure 5.

### 2.5 The test-positive proportion

The number of symptomatic and asymptomatic cases are linearly dependent on the symptomatic and asymptomatic testing probabilities respectively. The probability of testing can change over time. If these changes are not known and accounted for, then estimates of infection incidence from these case time series will be biased. If the total number of tests are known, then the proportion of tests that are positive (commonly referred to as the test-positive proportion) can be calculated, which is a surveillance indicator commonly used to monitor infection incidence. The test-positive proportion of symptomatic tests, and the test-positive proportion of asymptomatic tests are both independent of their respective testing probabilities (Box 3). However, the total proportion of all tests conducted (symptomatic and asymptomatic) that are positive, is generally not independent of the testing probabilities. Notable exceptions are when either testing probability is zero, or when the testing probabilities are equal (random sampling, see Section 3.1).

The test-positive proportion of symptomatic tests is highly dependent on the background rate of symptoms, *S*(*t*) (Figure 7). When *S*(*t*) is zero the test-positive proportion is approximately constant, with small fluctuations around this constant reflecting the effective sensitivity of the test (when sampling is performed at a rate dependent on the probability of exhibiting symptoms, *F* (*τ*)). The value at which the test-positive proportion is (approximately) constant is independent of the (average) infection incidence. When *S*(*t*) is greater than zero, the test-positive proportion decreases, and fluctuations in the test-positive proportion due to fluctuations in *I*(*t*) become more pronounced. For the extreme case in which *S*(*t*) is one, the test-positive proportion would be equal to the infection prevalence — this is equivalent to all testing being random (as symptoms no longer indicate an increased probability of infection) — but this is not a likely scenario. In general, *S*(*t*) will vary over time — potentially due to the circulation of other pathogens with similar symptom profiles — and so the effect *S*(*t*) has on the test-positive proportion of symptomatic tests will also vary over time making inference of the infection incidence difficult when estimates of *S*(*t*) are unavailable. In Figure 8 we demonstrate the potential effect of a time-varying value of *S*(*t*) on standard influenza surveillance — which relies on syndromic surveillance of those who exhibit influenza-like illness (ILI) — due to the co-circulation of SARS-CoV-2 (which also causes the symptoms consistent with ILI).

**Figure 7.**
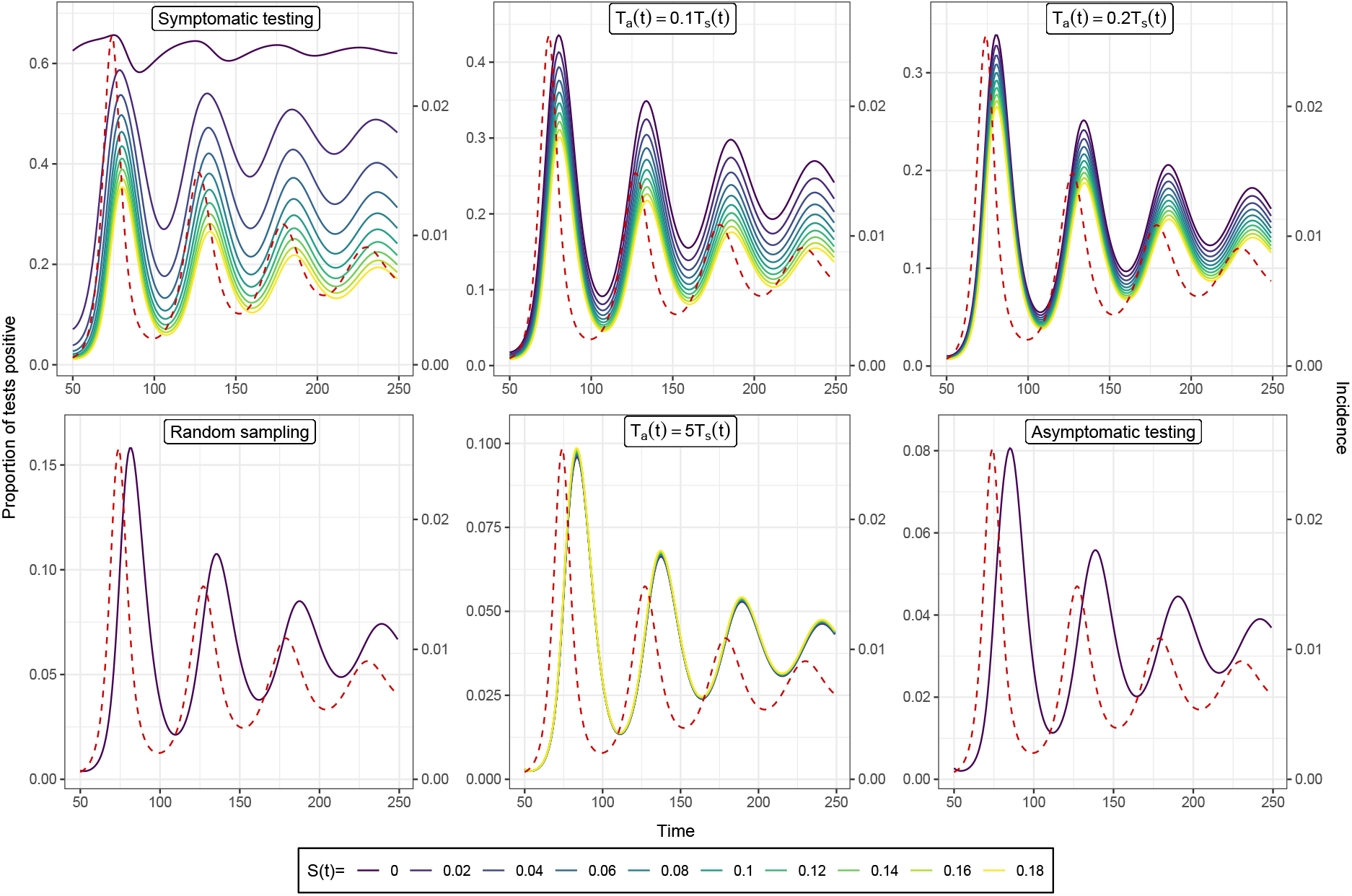
The effect of *S*(*t*) on the time series for the test-positive proportion. The time series of the test-positive proportion for six different testing regimes (with different symptomatic and asymptomatic testing probabilities): symptomatic testing (*T*_*a*_(*t*) = 0); *T*_*a*_(*t*) = 0.1*T*_*s*_(*t*); *T*_*a*_(*t*) = 0.2*T*_*s*_(*t*); random testing (*T*_*a*_(*t*) = *T*_*s*_(*t*)); *T*_*a*_(*t*) = 5*T*_*s*_(*t*); and asymptomatic testing (*T*_*s*_(*t*) = 0). The time series for the test-positive proportion were calculated assuming different constant values of *S*(*t*) (coloured lines) for a single example time series of infection incidence (red dashed line, right y-axis) which was simulated using an SIRS model [18]. The distribution of *F* (*τ*) was composed of *f*_1_(*τ*) and *f*_2_(*θ*) (see Section 2.1). *f*_1_(*τ*) was assumed to be gamma distributed with a mean of 3 days (shape parameter = 9, rate parameter = 3). *f*_2_(*θ*) was assumed to be one minus the cumulative distribution function of a gamma distributionwith a mean of 8 days (shape parameter = 40, rate parameter = 5). The distribution of *P* (*τ*) was chosen to match an empirical distribution for SARS-CoV-2 PCR positivity [17], with *ϵ* assumed to be zero.

**Figure 8.**
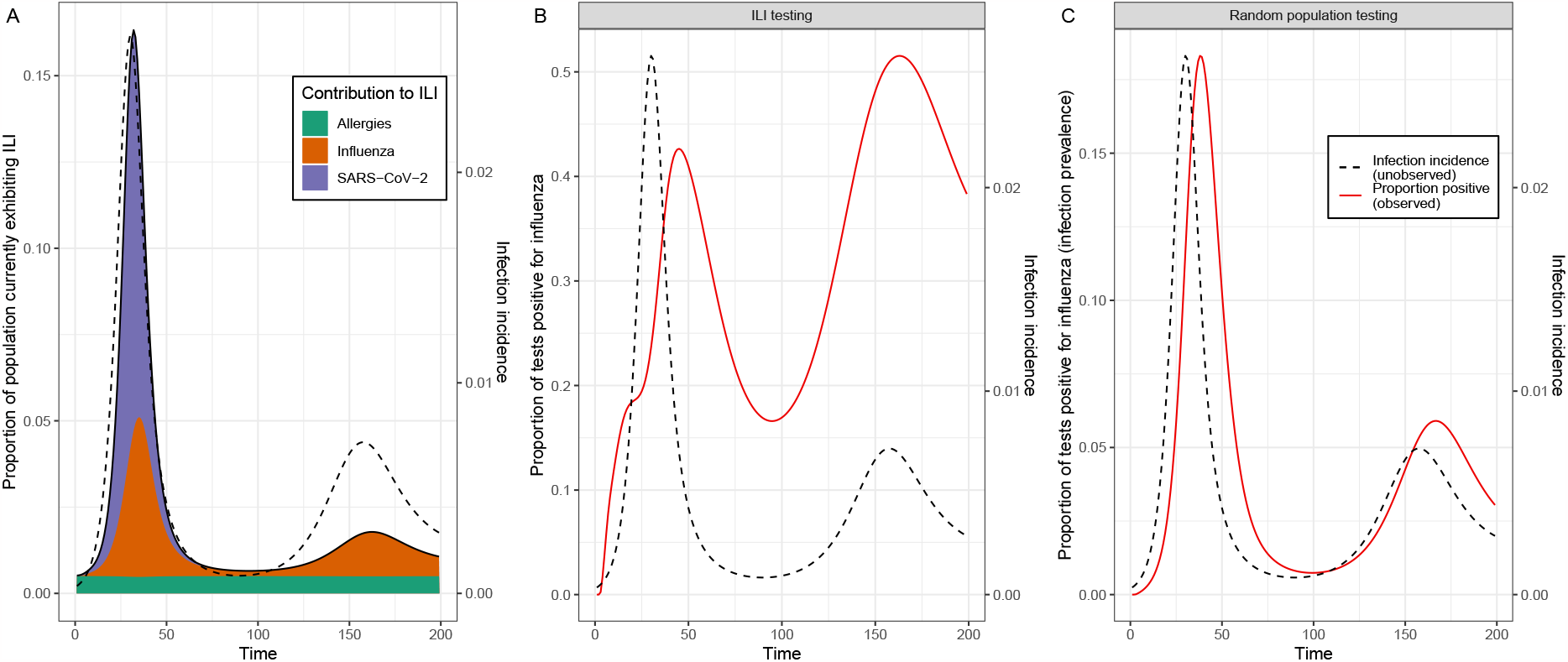
Example of the effect of a time varying value of *S*(*t*) on the test-positive proportion of influenza surveillance indicators. (A) The proportion of the population exhibiting influenza-like illness (ILI) (symptomatic with fever and cough) (left axis). This is a combination of those exhibiting ILI due to infection with influenza (orange), and the background proportion symptomatic *S*(*t*) (purple and green). The value of *S*(*t*) is the combination of a constant component due to a constant proportion of the population exhibiting allergies (with symptoms that match ILI definition) (green) and a time-varying component due to (for example) an epidemic of SARS-CoV-2,which can also cause the symptoms consistent with ILI (simulated using an SIR model [18], with individuals symptomatic for four days following infection) (purple). Also shown is the infection incidence for influenza (the pathogen of concern) (black dashed line, right y-axis) simulated using an SIRS model [18]. (B, C) The observed test-positive proportion (red line) shown for (B) ILI (symptomatic) testing and (C) random population testing (infection prevalence). Note that the time series of the test-positive proportion for random population testing is independent of *S*(*t*), but still does not match up perfectly with infection incidence (black dashed line, right y-axis). The time series of the test-positive proportion for ILI (symptomatic) testing is highly dependent on *S*(*t*) and we see that its second peak is greater than the first, despite the second peak for infection incidence being far lower than the first. The distribution of *F* (*τ*) used was composed of *f*_1_(*τ*) and *f*_2_(*θ*) (see Section 2.1). *f*_1_(*τ*) was assumed to be gamma distributed with a mean of 3 days (shape parameter = 9, rate parameter = 3). *f*_2_(*θ*) was assumed to be one minus the cumulative distribution function of a gamma distribution with a mean of 4 days (shape parameter = 40, rate parameter = 10). The distribution of *P* (*τ*) was chosen to match an empirical distribution for SARS-CoV-2 PCR positivity [17], with *ϵ* assumed to be zero. The distribution of *P* (*τ*) takes the form *logit*^*−*1^ (*β*_1_ + *β*_2_(*τ − C*) + *δ*_*τ−C>*0_ *× β*_2_*β*_3_(*τ − C*)) where *δ*_*τ−C>*0_ is 1 if *τ − C >* 0 and 0 if not [17]. The parameters used were: *β*_0_ = 1.51, *β*_1_ = 2.19, *β*_2_ = *−*1.1 and *C* = 3.18.

#### Box 3

**The proportion testing positive**

**Symptomatic testing**

The proportion of symptomatic tests that return a positive result can be written by dividing equation 8 by the symptomatic testing proportion, *T*_*s*_(*t*)*n*_*s*_(*t*), where *n*_*s*_(*t*) is given by equation 3. This gives:

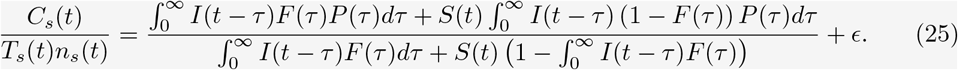

When *S*(*t*) = 1 this equation is identical to that of random sampling (equation 11) as now all individuals have the same probability of testing. When *S*(*t*) = 0 the equation takes the form:

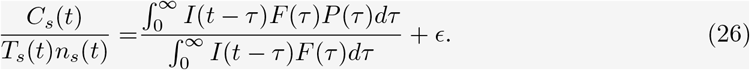

In general the value of the expression will vary over time (depending on the relative distributions of *F* (*τ*) and *P* (*τ*)), but fluctuations will be small and only reflect the relative asymmetry of the numerator and denominator. When *I*(*t*) is constant this equation gives the effective sensitivity for a sampling distribution describing the presence of symptoms, *F* (*τ*) (see equation 12).

**Asymptomatic testing**

The proportion of asymptomatic tests that return a positive result can be written by dividing equation 9 by the asymptomatic testing proportion, *T*_*a*_(*t*)*n*_*a*_(*t*), where *n*_*a*_(*t*) is given by equation 27. This gives:

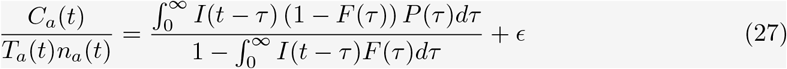

which has no dependence on *S*(*t*). The value of 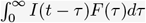 will usually be small and so the denominator will only mildly influence the value of the expression.

**All testing**

The overall proportion of tests that return a positive result can be written in terms of the total cases, and the total proportion tested.

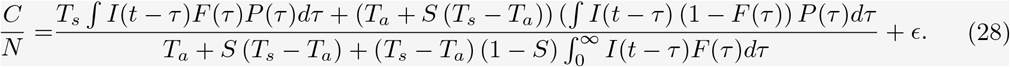

This expression is now dependent on both *S*(*t*) and the symptomatic and asymptomatic testing probabilities, *T*_*s*_(*t*) and *T*_*a*_(*t*). When *T*_*s*_(*t*) = *T*_*a*_(*t*) the equation is identical to that of random sampling (equation 11) and similarly for when *S*(*t*) = 1. As expected, when *T*_*s*_(*t*) = 0 we obtain equation 27 and when *T*_*a*_(*t*) = 0 we obtain equation 25.

Similar to the test-positive proportion of tests obtained using random sampling (infection prevalence, see Section 3.1), the test-positive proportion of asymptomatic tests is independent of *S*(*t*). Although the test-positive proportion of asymptomatic tests is not equal to the infection prevalence, its time series does have a similar mathematical relationship with infection incidence. The relationship between the test-positive proportion of asymptomatic tests and the infection incidence is dependent on the distributions *F* (*τ*) and *P* (*τ*), with the exact mathematical description provided in Box 3. When considering the test-positive proportion of all tests (symptomatic and asymptomatic), the resulting time series is dependent on the relative values of the symptomatic and asymptomatic testing probabilities, and the value of *S*(*t*) (Figure 7) — and of course how they change over time. The dependence on *S*(*t*) is most pronounced when the symptomatic testing probability is much larger than the asymptomatic testing probability.

### 3.6 Imperfect test specificity

When test specificity is imperfect (*ϵ >* 0), a constant proportion (*ϵ*) of all tests will be positive irrespective of infection status. Thus, imperfect test specificity will affect any surveillance indicator that is dependent on testing (e.g., all case time series as defined in this article). The number of cases (asymptomatic or symptomatic) identified due to imperfect test specificity will depend (linearly) on the number of tests administered (which can vary over time) as shown by equations 8 and 9. If one instead considers the proportion of tests that are positive (cases divided by the number of tests), then imperfect test specificity introduces a constant term (with the value given by *ϵ*) (Box 3) that will bias any case time series upwards. The magnitude of this bias depends on the test-positive proportion. The lower the test-positive proportion, the larger the relative value of this constant term, and the larger the upward bias in the number of cases. In general, the test-positive proportion will vary over time and so the impact of this bias will vary over time.

The test-positive proportion is highest when only symptomatic testing is conducted, and lowest when only asymptomatic testing is performed (Figure 7). Consequently, the greater the relative value of the symptomatic testing probability (relative to the asymptomatic testing probability), the higher the test-positive proportion of all tests (symptomatic and asymptomatic). When all testing is symptomatic, the higher the rate of background symptoms, *S*(*t*), the lower the test-positive proportion. When all testing is asymptomatic, the test-positive proportion is not dependent on *S*(*t*). The test-positive proportion is also dependent on the infection incidence, in general the lower the infection incidence, the lower the test-positive proportion. In the special case in which all testing is symptomatic and there are no background symptoms (*S*(*t*) = 0), then the test-positive proportion will not depend on the average rate of infection incidence, but will fluctuate around a constant value (effective sensitivity, see Box 3) due to variation in the rate of infection incidence (Figure 7A).

## 4 Considerations for future surveillance

### 4.1 Understanding the data-generating process

We have mathematically described the process by which case-based surveillance generates data and how these data are related to the time series of infection incidence. Accurate estimates of the infection incidence are required for a range of analyses that support disease management including estimation of the time-varying effective reproduction number, forecasting, and intervention scenario analysis [20, 21]. In theory, if the data-generating process is understood, and the individual processes (e.g., testing behaviour, probability of testing positive following infection) on which the data-generating process depends are well characterised, then the infection incidence can be estimated [21]. However, most of the individual processes are rarely known, or may have substantial uncertainty associated with them. Data-generating processes that depend on fewer individual processes are thus often best for analysis, as the analysis will likely include fewer biases, and less uncertainty. We have demonstrated that infection prevalence, measured using random sampling, is dependent on relatively fewer processes than other traditional forms of case-based surveillance — depending only on the probability of testing positive as a function of time since infection. Infection prevalence studies were performed during the COVID-19 pandemic [22, 10], which enabled robust estimates of the infection incidence [23], infection fatality ratio, infection hospitalisation ratio, and case ascertainment rate [24].

Routine surveillance indicators (e.g., case incidence, test-positive proportion, ILI etc) are often assumed to be reasonable correlates for infection incidence. Our analysis, which uses a mathematical description of the processes that generated the data for these indicators, demonstrates that this assumption is not reasonable and estimates of infection incidence based on such data could be heavily biased. For example, the Australian National Disease Surveillance Plan for COVID-19 lists the “Proportion of PCR tests conducted that are positive” as a key indicator for monitoring trends in infection and immunity [25]. However, we have shown that in general, the time series is dependent on the relative testing rates of symptomatic and asymptomatic individuals. Even when the vast majority of testing is symptomatic —which is very likely for routine surveillance (e.g., for influenza) — and so there is no dependence on testing rates, we have shown that the test-positive proportion of symptomatic tests is highly dependent on the background symptom rate, *S*(*t*). The background symptom rate can change over time, particularly if there is another circulating pathogen that causes a similar set of symptoms. Inference based on any such time series not accounting for *S*(*t*) would likely be highly biased with inaccurate estimates of the infection incidence and how it changes over time. Even when there is no background symptom rate (*S*(*t*) = 0), we have shown that the test-positive proportion of symptomatic tests does not reflect the infection incidence.

### 4.2 A holistic approach for surveillance design

Integration of multiple sources of data can improve estimates of infection incidence and downstream analyses that rely on infection incidence. In general, with data collected describing one epidemic time series (e.g., total daily cases) the infection incidence can only be inferred if the entire data generating process has been quantified. If multiple complimentary data sets are collected (e.g., total daily cases, and data describing test-seeking behaviour) then the infection incidence can still be inferred even if some individual processes, which compose the data-generating process, are not quantified [26]. When designing the surveillance system for a pathogen, it is thus necessary to consider what epidemiological insights can be gained by jointly analysing all available data, not just individual data sets. Some data may seem unimportant when considered in isolation, but may be crucial to the robust analysis of another data set(s).

We have demonstrated that the proportion of the population exhibiting symptoms is highly affected by the background symptom rate, and so individually a surveillance system based on quantifying the proportion of the population that is symptomatic (i.e., syndromic surveillance or symptom surveys) would be ineffective for quantifying infection incidence if other pathogens with similar symptom profiles were circulating (e.g., SARS-CoV-2, influenza and respiratory syncytial virus have similar symptom profiles). However, if combined with another epidemic time series that is also affected by *S*(*t*) (though in a different way) it might allow the dynamics of *S*(*t*) to be quantified and accounted for. Additionally, where symptom-based surveillance also collects data on whether an individual sought a test [27], then test seeking probabilities can be inferred — this information is useful for inference using case-based surveillance data which are dependent on these test seeking probabilities.

### 4.3 Balancing the different objectives of surveillance

In some circumstances there will be multiple competing objectives for case-based surveillance. The relative importance of these objectives will be resource and context specific. For example, consider two possible objectives for identifying cases: (1) to reduce ongoing transmission by isolating infected individuals, and quarantining their recent contacts [28], and (2) to quantify how infection incidence is changing over time. Using random sampling to measure infection prevalence might be the optimum choice (see Section 3.1) for surveillance when the second objective is a priority. However, if instead the first objective was the priority, surveillance strategies that return a higher proportion of positive tests (e.g., symptomatic testing) might be preferred [29]. This would allow a greater proportion of infections to be identified from a possibly limited supply of tests. In reality, the optimum solution is likely to contribute resources to both surveillance strategies, with resources weighted based on the relative importance of the objectives, and analysis should be performed using the appropriate data stream.

When a test has imperfect specificity, a constant proportion of all tests will be positive. The test-positive proportion is thus not expected to go to zero, and will only reach a minimum value defined by one minus the specificity. The lower the test-positive proportion is, the lower the power of a positive test to correctly predict infection. We have shown that the test-positive proportion is low when the infection incidence is low, except for symptomatic testing when there is a low rate of background symptoms in the population. Therefore, in certain situations when it is important that positive cases be accurately identified [30], symptomatic testing with strict symptom definitions (symptoms less likely to appear in the uninfected population) might be preferred. Optimum surveillance strategies will also depend on the timing of the increased probability of testing positive following infection. For example, if the timing is short and specific, it may be necessary to identify symptomatic features (e.g., onset of a specific symptom) where the timing closely aligns with the increased probability of testing positive. If instead infected individuals remain positive for a long period, then testing does not need to be so targeted and random sampling might be of greater utility.

### 4.4 Extensions of the mathematical description

We have presented a relatively simple model for the case-based surveillance of a pathogen. In our model individuals test with different probabilities based on their (binary) symptom status. The model could readily be adapted to more specific situations in which there is more knowledge available on test-seeking behaviours. For example, including a time-varying testing probability based on the time since symptom onset, or including testing probabilities that vary based on age or the severity of infection. The mathematical description is also easily adaptable to other forms of data collection. For example, serological studies might be modelled using a different distribution of *P* (*τ*), in which the probability of testing positive remains high for a much longer period, or surveillance based on severe outcomes might instead describe the probability of hospital admission or death (as a function of time since infection). If the data-generating processes of multiple epidemic time series can be defined, then all time series can be used simultaneously to estimate the infection incidence; this would allow the uncertainty present in any individual process to be minimised.

## 5 Conclusions

Designing an effective surveillance strategy is complicated [31], with the optimal strategy likely depending on the pathogen, setting, and resource constraints. We have developed a general mathematical description for the data generating process of case-based surveillance. The description highlights the complex relationships between the time series of surveillance indicators and the time series of infection incidence, an important quantity for informing public health decision-making. Analysis of surveillance time series should be cognisant of these complex relationships, and not assume that the time series is a reasonable proxy for infection incidence. By considering the full mathematical description of such data generating processes, future surveillance strategies could be designed to improve estimates of the infection incidence by minimising potential sources of uncertainty and bias.

## Data Availability

This is a simulation study and so there is no data.

## Supplementary figures

**Figure S1:**
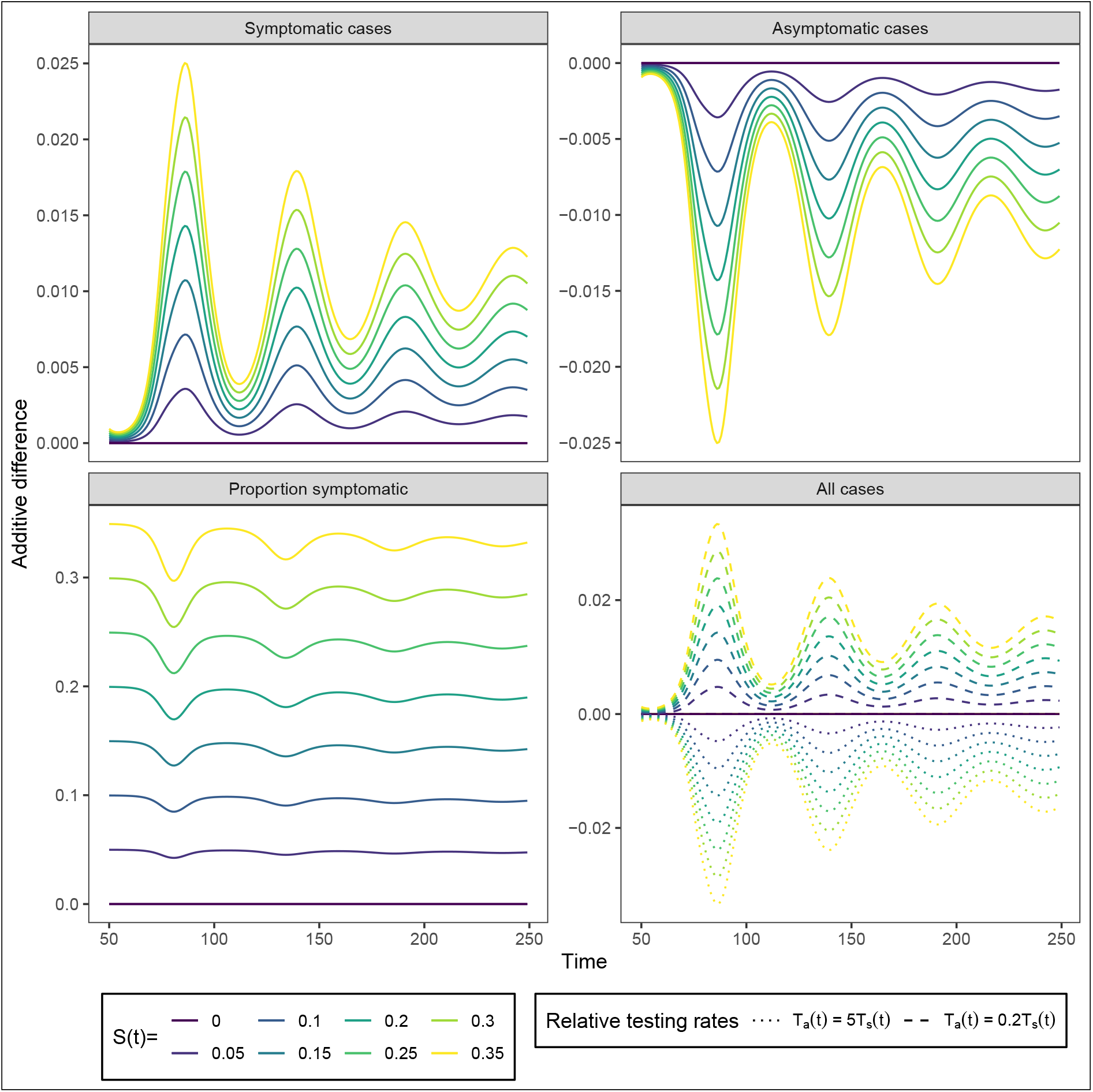
The additive effect of *S*(*t*) on surveillance indicators. The additive difference between the time series of a surveillance indicator when *S*(*t*) = 0 and the time series of the same surveillance indicator when *S*(*t*) *>* 0 (coloured lines). The time series of additive differences are shown for four possible surveillance indicators: symptomatic cases; asymptomatic cases; proportion of the population symptomatic; and cases identified through mass testing (combined total of symptomatic and asymptomatic cases). The time series for all surveillance indicators and the time series of infection incidence are shown in Figure 5. The additive difference between the time series for cases identified through mass testing were shown for different relative testing rates: *T*_*a*_(*t*) = *T*_*s*_(*t*) (random testing); *T*_*a*_(*t*) = 5*T*_*s*_(*t*) (higher testing probability for asymptomatic individuals); and *T*_*a*_(*t*) = 0.2*T*_*s*_(*t*) (higher testing probability for symptomatic individuals). Note that for random testing the additive difference was zero for all values of *S*(*t*).

**Figure S2:**
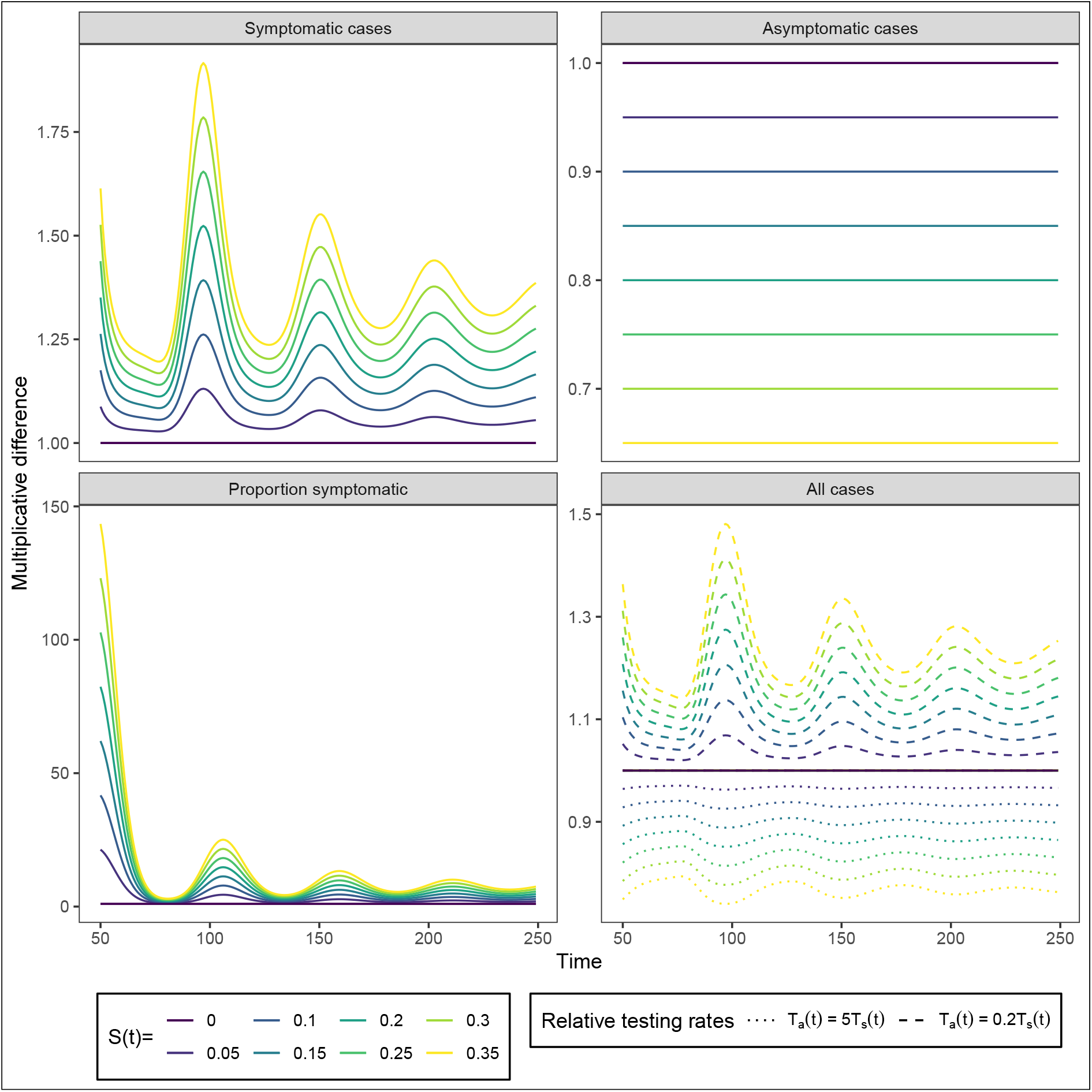
The multiplicative effect of *S*(*t*) on surveillance indicators. The multiplicative difference between the time series of a surveillance indicator when *S*(*t*) = 0 and the time series of the same surveillance indicator when *S*(*t*) *>* 0 (coloured lines). The time series of multiplicative differences are shown for four possible surveillance indicators: symptomatic cases; asymptomatic cases; proportion of the population symptomatic; and cases identified through mass testing (combined total of symptomatic and asymptomatic cases). The time series for all surveillance indicators and the time series of infection incidence are shown in Figure 5. The multiplicative difference between the time series for cases identified through mass testing were shown for different relative testing rates: *T*_*a*_(*t*) = *T*_*s*_(*t*) (random testing); *T*_*a*_(*t*) = 5*T*_*s*_(*t*) (higher testing probability for asymptomatic individuals); and *T*_*a*_(*t*) = 0.2*T*_*s*_(*t*) (higher testing probability for symptomatic individuals). Note that for random testing the multiplicative difference was one (no difference) for all values of *S*(*t*).

